# Amputation in Kingdom of Saudi Arabia: Etiology, Characteristics and Clinical status from a Large Tertiary Rehabilitation Center

**DOI:** 10.1101/2022.03.09.22272172

**Authors:** Enas M. Shahine, Mohamed T. Silarbi, Abdullah Alzeer, Mostafa Bukhamseen, Khalid Mohammed Alzaraa, Iram Saba

## Abstract

**Introduction:** Limb amputation significantly impacts the patient’s physical, emotional, and social life. Multiple etiological factors may lead to limb amputation. Unfortunately, there is limited data about amputation in Saudi Arabia. Since Sultan Bin Abdulaziz Humanitarian City (SBAHC) is a tertiary centre receiving such patients, we collected data from ten years to look at the etiology, characteristics, and clinical impact of amputations in the Kingdom.

**Methods:** A retrospective study of 1409 amputee patients’ data at SBAHC collected over ten years included demographic variables, etiology, site, level, and type of amputation. We also collected Functional independent measurement (FIM) scores for 618 patients.

**Result:** Males constitute the majority of amputees (75.7%) and the average age in males was higher in males compared to females (45 vs 36 years respectively, p<0.001). Vascular diseases (42.3%) and Trauma (36.8%) were the leading cause of amputation in this cohort. Diabetes mellitus was the most frequent comorbidity (40.5%), followed by hypertension (26.2%). Transtibial amputations were the most typical (22.58%), followed by trans-femoral amputation (14.12%). Traumatic trans-femoral amputation was more prevalent among young adults than traumatic trans-tibial amputation. Trauma-related amputation cases were highest in the age group of 21-30 years (69.2%), while vascular related amputations were highest in the age group of 70 and above years (89.5%). FIM scores improved significantly in locomotion (33.6%) followed by transfer (30.6%) and self-care (16.4%) at six-month post discharge compared to admission (all p-values<0.01).

**Conclusion:** Vascular pathology arising due to chronic diseases is the primary risk factor that may lead to amputation and warrant primary and secondary prevention programs. Seatbelt enforcement can significantly decrease amputation related to Road Traffic Accidents, which has been the second most familiar cause of amputation.

## Introduction

Amputation has a profound and long-lasting impact on an individual’s economic, psychological, and social status, and the prevalence of amputations performed has been rising worldwide [1]. Lower limb loss was estimated to affect 1.6 million people in the United States in 2005, with that figure expected to more than double to 3.6 million by 2050 [2]. Complications arising from various peripheral vascular diseases and/or diabetes form the most important causes of amputations. However, extremity traumas and, to a lesser extent, cancer and other etiologies also contribute [3]. Studies have shown that age and gender, and etiology play a significant role in the pattern of limb loss. Limb loss is more common in people with comorbidities, young and men [4]. Most patients requiring prosthetic restoration and rehabilitation services are older adults who have had lower limb amputations due to vascular disease [5]. The lifetime risk of amputation in the elderly is 10–15%, 10–30 times higher than in the general population. The data also shows that Trauma and cancer-related amputations are decreasing, while congenital amputations remain stable. Studies show that diabetes is responsible for at least half of non-traumatic lower-limb amputations worldwide [6]. The studies regarding amputation among countries in Asia have many similarities, as trauma and disease incidence occur in similar patterns. In contrast, the patterns differ from those in the United States, Denmark, Finland, and Australia [7], where peripheral vascular diseases, particularly arteriosclerosis, are the most common cause of amputation.

The incidence rate of peripheral vascular diseases, diabetes, and road traffic accident-toll in Saudi Arabia are also on the rise [8, 9]. Consequently, it may also have resulted in the rising cases of amputations in this country. More data needs to be published to assess better the etiology, characteristics, and risk factors for amputation in this country. However, there is a lack of a National Registry for Amputations. This fact has increased the interest in studying the impact of etiology, characteristics, and clinical status on amputees. Sultan bin Abdul Aziz Humanitarian City (SBAHC), a tertiary care rehabilitation centre, adopted proper evaluation management and quality of life assessment for its clients where we conducted this study. Besides, identifying the etiology, characteristics, and risk factors for amputation will assist health care providers and patients in rehabilitation sessions in planning and establishing effective and timely interventions. The present study’s purpose may also help improve rehabilitation services so that amputee patients can be functionally independent and productive members of their communities.

## Materials and Methods

This investigation is a retrospective hospital-based study with 1409 amputee patients at Sultan bin Abdul Aziz Humanitarian City (SBAHC), Riyadh, Saudi Arabia, from June 2010 to June 2020. All amputees’ medical records were evaluated using a pre-designed case report form to determine the pattern of amputations and etiological factors. Inclusion criteria included all patients with amputations who visited outpatient limb loss clinics. The exclusion criteria included the patients whose medical records were incomplete during the study period regardless of the level, site or type of amputation. Collected data included demographic variables, etiology, site, level, and type of amputations. In addition, the cohort distribution from different provinces, namely Central, Northern, Southern, Eastern, and Western, was collected in addition to patient nationality.

Out of the selected cohort, 618 patients were required to be admitted because of pre- and post-prosthetic rehabilitation. This cohort was used to assess functional independence using a functional independent measurement score (FIM Score) at admission and six months after discharge. Clinical evaluation in weight, height, BMI, and systolic/diastolic blood pressure was also collected from the patient’s medical files. Progress with rehabilitation, if any, was recorded during the follow-up. The FIM instrument contains 18 items classified into six major scales: self-care, sphincter control, transfer, locomotion, communication, and social cognition. On each item, the patients rated their level of independence using a 1–7 scale, on which 1 means assistance was needed, and 7 means a person was entirely independent.

### Statistical Analysis

The Pearson chi-square test was used to determine any significant relationship between the measured variables. The data were presented as the mean, standard deviation for continuous normal variables, and for continuous non-normal variables, the data were presented as median (Q1-Q3). The relationship between etiology, age, and level of amputation was calculated using the pivot graphs and tables. A P-value of 0.05 was considered significant. Each case was interviewed during admission and discharge using a self-reported version of the FIM instrument. Summing up responses to each item gave us a scale score ranging from several items to seven times the number of items, i.e., the total FIM score ranges from 18 to 126. FIM improvement (%) was calculated by subtracting FIM at admission from FIM at discharge, and this score served as an outcome measure in the present study.

## Results

The demographic characteristics of the study subjects were presented in table 1. A total of 1409 patients were recruited, out of which 75.6% were male and 24.4% were female. The patients’ ages varied between 1 and 96 years, with a mean age of 45 years for males and 36 years for females. Out of the total population studied, Saudi nationals constituted 86.9%, and the rest were non-Saudis (13.1%). Most of the studied population (60.3%) was from the Central region of Saudi Arabia, followed by the Western (16.7%), Southern (11.4%), Eastern (7.5%) and Northern (4.2%) regions. Most amputees were affected by vascular and traumatic amputations (42.4% and 36.9%, respectively), followed by congenital (14.4%) and infection/others (6.3%). The study cohort majorly consists of patients with a duration of amputation of 5 years (40.4%), followed by 6–10 years (29.7%), 11–15 years (13.2%), > 20 years (10.8%), and then 16–20 years (6.0%). Among comorbidities, the most prevalent was diabetes (40.5%), followed by hypertension (26.3%), cardiac diseases (8.4%) and hyperlipidemia (5.3%).

**Table 1.** Demographic Characteristics of study subjects

| Total |  | 1409 | Male (1066) | Female (343) | p-value |
| --- | --- | --- | --- | --- | --- |
| Age Mean (SD) |  | 43 (21) | 45 (20.8) | 36 (20.0) | < 0.001 |
| Nationality N (%) | Saudi | 1224 (86.8) | 920 (75.1) | 304 (24.9) | < 0.001 |
|  | Non-Saudi | 185 (13.1) | 146 (78.9) | 39 (21.1) | < 0.001 |
| Province N (%) | Central | 849 (60.2) | 650 (76.5) | 199 (23.5) | < 0.001 |
|  | Northern | 59 (4.1) | 37 (62.7) | 22 (37.3) | 0.006 |
|  | Southern | 160 (11.3) | 126 (78.7) | 34 (21.3) | < 0.001 |
|  | Western | 235 (16.6) | 181 (77.1) | 54 (22.9) | < 0.001 |
|  | Eastern | 106 (7.5) | 72 (67.9) | 34 (32.1) | < 0.001 |
| Etiology N (%) | Trauma | 519 (36.8) | 425 (81.8) | 94 (18.2) | < 0.001 |
|  | Vascular | 597 (42.3) | 479 (80.2) | 118 (19.8) | < 0.001 |
|  | Congenital | 204 (14.4) | 109 (53.4) | 95 (46.6) | 0.163 |
|  | Infection | 89 (6.3) | 53 (59.5) | 36 (40.5) | 0.01 |
| Duration of amputation N (%) | ≤5 | 566 (40.3) | 447 (78.9) | 119 (21.1) | <0.001 |
|  | 6 - 10 | 417 (29.6) | 329 (78.8) | 88 (21.2) | < 0.001 |
|  | 11-15 | 186 (13.2) | 137 (73.6) | 49 (26.4) | < 0.001 |
|  | 16 - 20 | 84 (5.9) | 55 (65.4) | 29 (34.6) | < 0.001 |
|  | >20 | 152 (10.7) | 95 (62.5) | 57 (37.5) | < 0.001 |
| Comorbidity N (%) | Diabetes | 570 (40.4) | 468 (82.1) | 102 (17.9) | < 0.001 |
|  | Hypertension | 370 (26.2) | 301 (81.5) | 69 (18.5) | < 0.001 |
|  | Hyperlipidemia | 74 (5.2) | 63 (85.1) | 11 (14.9) | < 0.001 |
|  | Cardiac | 118 (8.3) | 102 (86.4) | 16 (13.6) | < 0.001 |
|  | Others | 277 (19.6) | 193 (69.6) | 84 (30.4) | < 0.001 |
| Weight Mean (SD) |  | 72.5 (32.4) | 72.4 (32.4) | 62.8 (31.7) | < 0.001 |
| Height Mean (SD) |  | 158.5 (54.6) | 158.50 ( 54.6) | 146.5 (48.3) | < 0.001 |
| BMI Mean (SD) |  | 30.2 (12.5) | 30.58 (12.5) | 26.9 (12.3) | 0.03 |
| Systolic Blood Pressure Mean (SD) |  | 125.9 (42.5) | 128.9 ( 42.5) | 120.6 (49.1) | < 0.001 |
| Diastolic Blood Pressure Mean (SD) |  | 76.3 (24.8) | 76.3 (24.8) | 74.5 (22.7) | 0.47 |

**Note:** Data is presented as prevalence (%). The difference between the groups is tested by Chi-square test. P<0.05 is considered significant.

When comparing the level of amputation with etiology, trans-tibial amputations were found to be the highest (22.6%), followed by trans-femoral amputations (14.1%) in the case of patients with vascular diseases. Among trauma patients, trans-femoral amputation was the highest (10.9%), followed by transtibial (9.6%). The amputation level among congenital cases was highest in the case of trans-radial (2.4%), followed by partial hand (1.8%) and trans-femoral (1.6%) (Figure 1).

**Figure 1.**
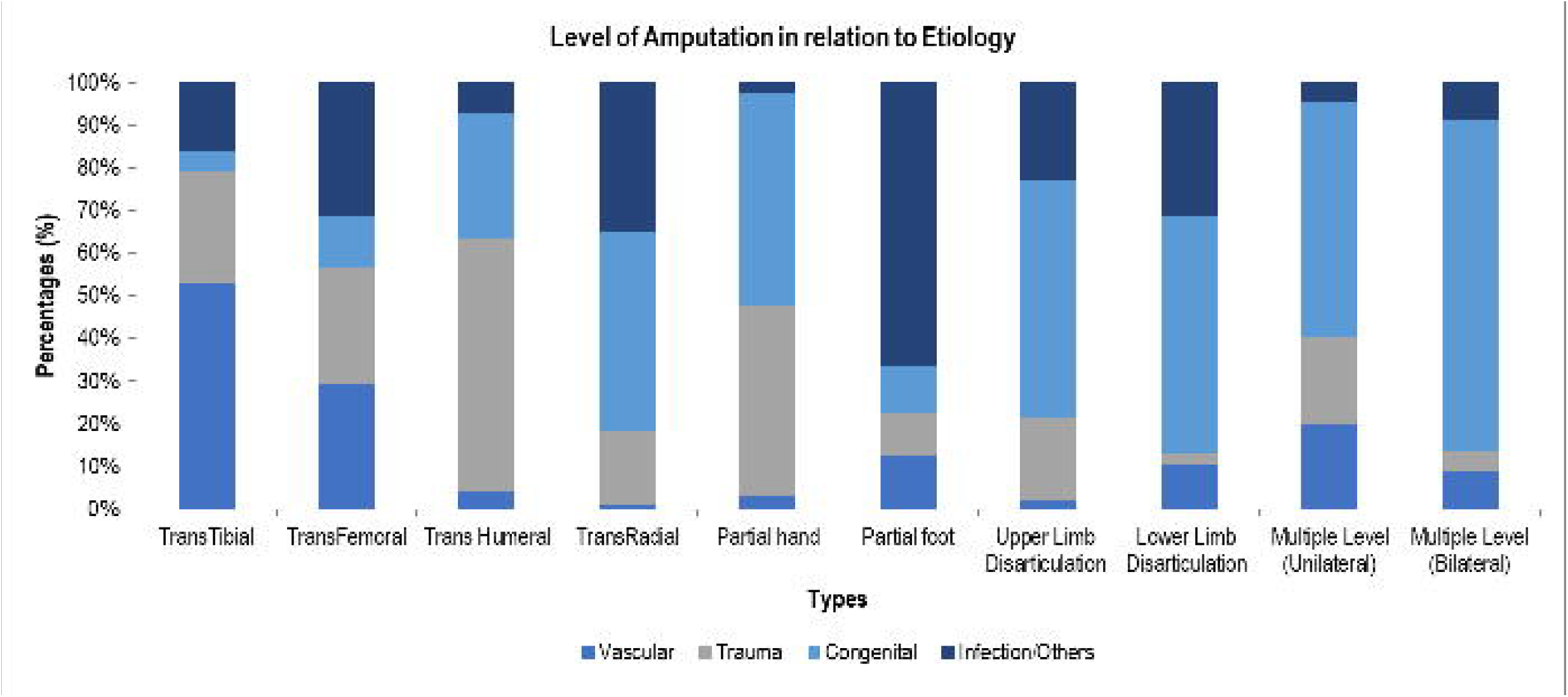
Level of amputation in relation to etiology.

When comparing the etiology of amputation with life span (Figure 2), we found that trauma-related amputation cases were highest in the age group between 21 to 30 years (69.2%) and lowest in people over 70 years of age (7%). The vascular related amputations were highest in the age group of 70 and above years (89.5%) and least in age-group of 0-10 years (3.9%). Congenital amputation were as expected most frequent in the age group 0–10 years (79.4%), followed by 11–20 years (37.5%), and was least in the age group 51–60 years (0.5%).

**Figure 2.**
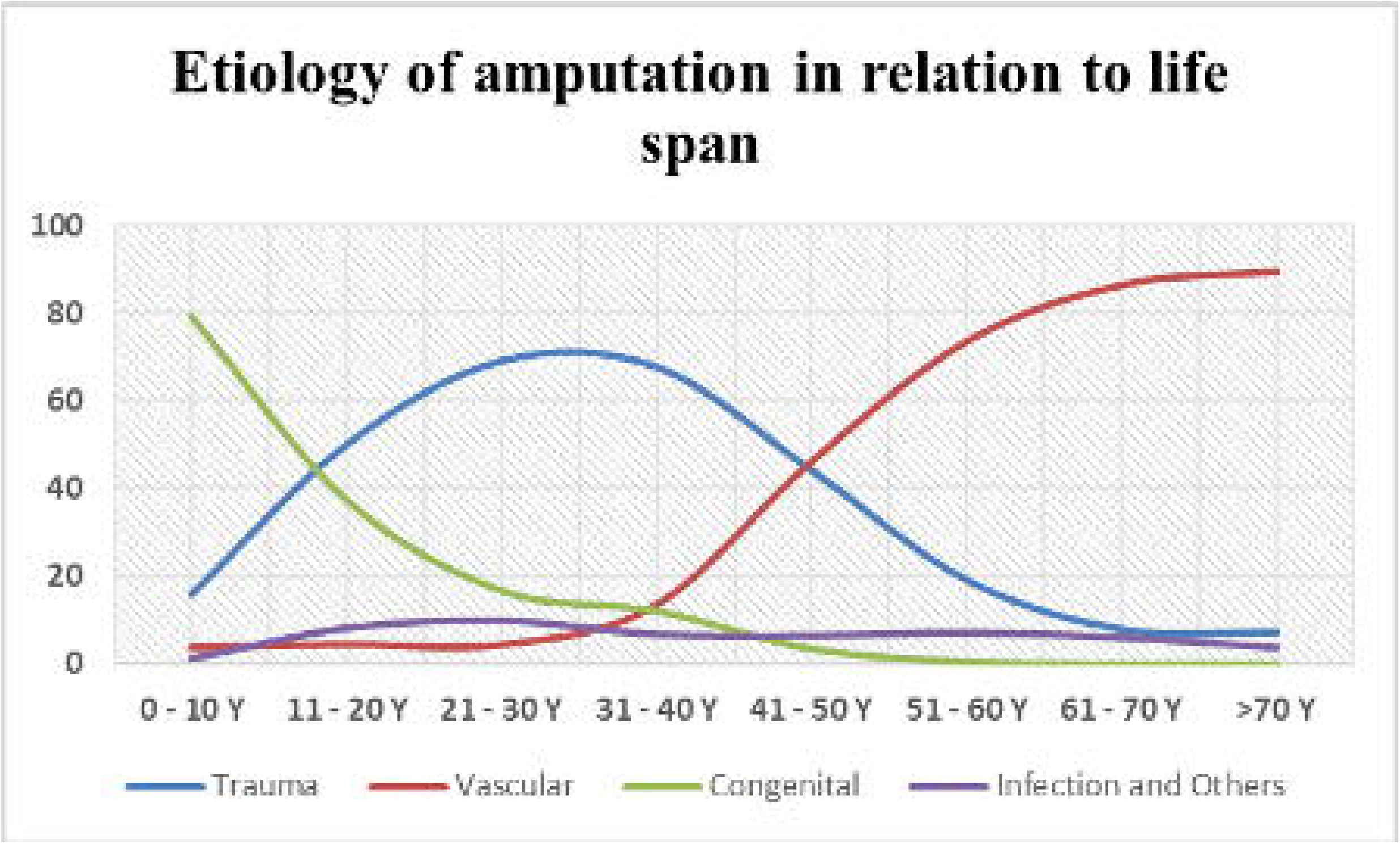
Etiology of amputation in relation to life span.

Admission and discharge FIM score data was presented in Table 2. There is a significant difference (p 0.005) between admission and discharge scores in self-care, transfer and locomotion (all p-values<0.001). Also, the overall FIM score improved 6-months post discharge as compared to the admission (p<0.001). FIM showed a maximum improvement in the case of locomotion (33.6%), followed by transfer (30.6%) and self-care (16.4%). An improvement of 15.1% was also found in the total FIM score.

**Table 2.** Patient’s functional status according to Functional Independence Measurement (FIM) Score value from admission to discharge

| FIM Scale<br>Mean (SD) | Admission | Discharge | P | Max<br>Score | Admission<br>(% score) | Discharge<br>(% score) | Improvement<br>(%) |
| --- | --- | --- | --- | --- | --- | --- | --- |
| Self-care | 29.6 (0.5) | 36.6 (0.3) | <0.001 | 42 | 71.4 | 87.8 | 16.4 |
| Sphincter-Control | 12.5( 0.2) | 12.5 (0.2) | 0.34 | 14 | 88.0 | 91.8 | 3.8 |
| Transfer | 11.0 (0.4) | 17.3 (0.2) | < 0.001 | 21 | 52.6 | 83.2 | 30.6 |
| Locomotion | 6.4 (0.3) | 10.8 (0.2) | < 0.001 | 14 | 45.9 | 79.5 | 33.6 |
| Communication | 13.6 (0.1) | 13.7 (0.1) | 0.19 | 14 | 97.6 | 98.8 | 1.2 |
| Social-Cognition | 20.3 (0.1) | 20.4 (0.1) | 0.29 | 21 | 97.0 | 97.9 | 0.9 |
| Total FIM | 89.8 (1.2) | 107.5 (1.1) | < 0.001 | 126 | 74.4 | 89.5 | 15.1 |

**Note:** Data is presented as prevalence (%). The difference between the groups is tested by Chi-square test. P<0.05 is considered significant.

## Discussion

Amputee incidence may differ not only from one country to another but also within the same country based on many factors: the degree of modernization, accessibility of medical and surgical care, and living standards. This study will give a better picture of the physical and pathological distribution of amputations in the five central provinces of Saudi Arabia. It is located in Riyadh, the capital of Saudi Arabia, so it is easily accessible to receive patients from the five major provinces of the Kingdom, namely: Central, Southern, Eastern, Western, and Northern Regions. This will be done to give a better picture of amputations’ physical and pathological distribution. This study aimed to present our experiences with major limb amputations and compare the results to those of similar studies conducted in other regions of the world to emphasize the differences in the pattern and indications for amputations and suggest relevant preventive interventions.

This study’s male predominance among amputees corresponds with other researchers that have found similar findings regardless of ethnicity and geographical distribution [10, 11]. This could be attributed to men having more severe peripheral artery diseases, higher smoking rates, and more susceptibility to road traffic accidents. On the other hand, estrogen and its effect on lowering vascular pathology may explain the lower incidence of amputation related to vascular disease [12]. The fact that diabetic patients have a 15-fold greater risk of amputation when coupled with the rising incidence of diabetes and vascular disease increases in lower limb amputation necessitates the importance of early detection, medical education, patient compliance, and reasonable glycemic control in this population.

Diabetic neuropathy that may lead to loss of sensation, abnormal gait, and deformity would increase the chance of foot pathology that increases the risk of lower limb amputation when coupled with vascular abnormality. In addition, the presence of chronic foot ulcers invites infection, especially deep wound and osteomyelitis infections that may end in amputation [13]. In a study from South Africa and India, infection and ulcers were the most common causes of amputation in diabetic individuals. At the same time, ischemia was the most common cause in nondiabetic people, which was not the case in our studied cohort. This could be related to the better care provided to this cohort since medical care is free and provided by secondary and tertiary care centres [14, 15].

In the studied cohort, Trauma is the second observed cause of amputation, which is similar to what has been found in developed countries, contrary to what has been found in developing countries, where Trauma is the first cause of amputation [16]. These variations in amputation patterns reflect differences in comorbidities. It has been demonstrated that the risk of amputation rises as the number of comorbidities grows, which is the finding of this study and could provide a scientific basis for the higher incidence when compared with Trauma. SBAHC, being a tertiary care centre, receives most of its patients within five years of amputation, reflecting the efficiency of this hospital in providing services for such patients. Although Trauma was ranked second in this study, it was the most common reason for amputation in young adults in their productive and active age group.

In this study, the relationship between etiology and amputation level demonstrates the significant variations between amputation and surgical decisions by medical staff. Most of our amputations were in the lower limbs, with transtibial amputation the most common regardless of the etiology [17]. This data supports previous findings that lower limbs are wounded more frequently than upper extremities, and diabetic gangrene is more common in the lower extremities than in any other part of the body. However, other studies have found that trans-femoral amputation is more common than transtibial, which could reflect either the worst lower limb infection that mandates trans-femoral amputation or surgical attitude [18, 19]. When comparing Trauma to the degree of amputation, the transfemoral and transtibial amputations rates were nearly the same, at 9.6% and 10.9%, respectively. That could be the effect of a wide age range in this cohort. According to research published in 1993, Trauma was the leading cause of lower limb amputation in Saudi Arabia (52.9%). The most common location of amputation was the trans-tibia, followed by the trans-femoral, trans-radial, partial hand, and trans-humeral [20]. Since then, there has been no data in PubMed about amputations in this country. This could be explained by the increasing incidence of comorbidities, especially diabetes, and the strengthening of safety belt rules in society [21].

Different age groups in our studied population showed variance in etiology. In our study, trauma-related amputations were most common in the age group of 20 to 40 years. This is consistent with a WHO report signalling that road traffic accidents are among the top causes of death in those aged 15 to 29 years old, as the first few months after receiving a license are extremely hazardous [22]. When considering the influence of age on amputations and its relationship to etiology, vascular disease is the most common cause. This was observed in the 3rd and 4th decades of life and was identical to the reports from other developed and developing countries’ studies [23].

Consequently, vascular disease-related amputation was most common in older people aged over 60 years. Furthermore, older people are more likely to have more than one ailment that may result in amputation due to vascular etiology [24]. Congenital limb deficiency, on the other hand, accounted for 79.4% of lower limb deficiency in children under the age of 10 years in this study, compared to 67% in the Krebs and Fishman study [25], 32% in the Yigiter et al. study [26], and 73.3% in the Boonstra et al. study [27]. This could be at the expense of trauma-related limb loss based on strengthening children’s car safety rules. In addition to data from tertiary centres receiving patients from all over the Kingdom, These demographic parameters of children with limb deficits showed a high degree of consistency throughout the literature.

There was a significant difference between FIM scores at admission and discharge and a total FIM gain during the hospitalization. On follow-up visits, we saw a considerable improvement in the total FIM score and its motor and transfer subscales. The average total FIM scores for admission and discharge were 89.811.2 and 107.51.1, respectively. An FIM score of greater than 108 indicates home independence [28]. On six-month follow-up visits, individuals in this study achieved this independence degree. This improvement was most noticeable in the self-care, transfer, and locomotion categories. The results showed an improvement in self-care, transfer, and locomotion in the FIM scores between admission and discharge, comparable with the findings of the Hall et al. study [29]. The research found that advances in the motor domain were much higher than gains in the cognitive domain, as the cognitive score was not the scope of this study, in addition to the fact that such patient categories are not associated with mental or cognitive defects.

This study’s strength comes from being conducted in the largest tertiary rehabilitation centre in the country, which was able to represent amputees nationwide and covers all age spectrums. The second strength is that amputees in this institution are subject to a multidisciplinary approach that would give better clinical, physical and mental assessment, as shown in this study. Finally, although this study was limited to being a retrospective study and receiving patients after five years, that would not provide data related to immediate amputation outcome.

## Conclusion

Vascular pathology caused by chronic illnesses is the key risk factor for amputation, necessitating primary and secondary preventive efforts. If patients and caregivers were better informed and instructed about post-amputation objectives, they would be more likely to adhere to professionally prescribed rehabilitation programs. A multidisciplinary team approach, in-depth understanding of the functional impacts of amputation, and a comprehensive and detailed examination of the patients and their surroundings should serve as the cornerstone for post-amputation follow-up programs. Seatbelt use can considerably reduce amputations caused by road traffic accidents, which are the second most common cause of amputation. This study may raise awareness among healthcare providers about the need for primary and secondary preventive strategies for limb loss.

## Data Availability

All data produced in the present study are available upon reasonable request to the authors

## Author Contributions

Conceptualization, E.M.S.; Methodology, M.T.S, A.Z., M.B.; Data curation, I.S.; Writing— original draft preparation, I.S.; Writing—review and editing, E.M.S., M.T.S., M.B.; Supervision, E.M.S., K.M.A. All authors have read and agreed to the published version of the manuscript.

## Acknowledgement

The authors would like to acknowledge the staff of the research center, nurses from the surgical department, and the department of health information system (HIS) at SBAHC.

## Institutional Review Board Statement

The study design and protocol was approved by the Ethics Committee for Scientific Research at SBAHC, Riyadh Saudi Arabia ,under the IRB No.03-2019-IRB with study code:01/SBACH/RH/2019.

## Informed Consent Statement

Informed consent was obtained from all subjects involved in the study.

